# Silent Manipulation of Mental Health Treatment Recommendations from a Large Language Model

**DOI:** 10.64898/2026.06.16.26355686

**Authors:** Roy H. Perlis

## Abstract

**Importance:** Large language models (LLMs) increasingly inform mental health decisions by patients and clinicians. Inference-time activation steering can shift model behavior on a target dimension without altering weights or prompts and without disclosure to users, allowing treatment recommendations to be silently changed for commercial or ideological reasons.

**Objective:** To determine whether directional activation steering can shift an open-weights LLM’s depression treatment recommendations.

**Design, Setting, and Participants:** This non-human subjects study applied directional activation steering to an open-weights LLM (DeepSeek V4 Flash) responding to 12 depression-advice scenarios (4 favoring medication, 4 favoring avoidance, 4 neutral), generated at 30 amplitudes from −1.5 to +1.5 in 0.1 increments plus an unsteered baseline.

**Exposures:** A single steering direction contrasting antidepressant medication with self-directed approaches (diet, exercise, meditation, dietary supplements), constructed from 16 paired training prompts and applied at the attention output of every transformer block; weights and system prompt were held constant.

**Main Outcomes and Measures:** The extent to which medication and four self-care categories were addressed, scored 0 to 3 by a human-validated LLM rater (Claude Opus 4.7), the medication-versus-self-care balance, and clinician referral, estimated per unit of amplitude using mixed-effects models with a scenario random intercept.

**Results:** Across 372 generations, steering produced a graded, dose-dependent shift in the medication-versus-self-care balance, which declined by 0.32 per unit of amplitude (β = −0.32; 95% CI, −0.39 to −0.25; P < .001); medication extent fell and self-care extent rose. The shift was largest for scenarios with no stated treatment preference (β = −0.44; 95% CI, −0.54 to −0.34; P < .001). A clinician referral appeared in 322 of 372 responses (87%) and did not vary with steering amplitude (P = .63).

**Conclusions and Relevance:** In this open-weights LLM providing depression treatment information, inference-time activation steering shifted treatment recommendations without altering weights, prompt structure, or safety outputs, with the largest effect among users expressing no treatment preference. These findings suggest a need for LLM disclosure standards and independent auditing as such models inform clinical decisions.

## Introduction

Large language models (LLMs) are increasingly applied to inform decision-making by health care consumers and clinicians.^1^ The behavior of these models is impacted by their initial weights as well as post-training interventions, including fine-tuning and system prompting that are typically not disclosed to end users. An alternative strategy, inference-time activation steering, directly alters model activations in order to shift behavior on a target dimension, while remaining undetectable by the user.^2,3^

These potential manipulations allow care recommendations to be silently altered for commercial or ideological reasons without disclosure.^4,5^ For example, in light of recent efforts to politicize antidepressant use, it is possible that activation steering could alter public health messages. To characterize this risk, I applied this method to an open-weights LLM positioned as a mental health information assistant and measured shifts in treatment options.

## Methods

This quality-improvement, non-human subjects study applied directional activation steering, via the open-source tool DS4,^6^ to an open-weights LLM (DeepSeek V4 Flash). Full model and output generation parameters are provided in the eMethods in Supplement 1.

A single steering direction contrasting antidepressant medication with self-directed approaches (diet, exercise, meditation, dietary supplements) was constructed from 16 paired training prompts and applied at the attention output of every transformer block; negative scale amplifies the medication direction and positive scale the self-directed direction (Supplement 1).^3^ Model weights and system prompt were identical across conditions.

Responses to 12 depression-treatment-advice scenarios, 4 preferring medication, 4 preferring to avoid, and 4 stating no preference (Supplement 1), were generated at 30 steering amplitudes from −1.5 to +1.5 in 0.1 increments, plus an unsteered baseline. Each response was scored by an LLM rater (Claude Opus 4.7) validated against a blinded human expert to examine extent to which medication, versus four self-care categories of diet, exercise, meditation, and nutrition, all on a 0 to 3 scale, were addressed. Association was estimated with linear mixed-effects models including a random intercept for scenario; whether the output included a clinician referral used a mixed-effects logistic model. Effects are reported per unit of steering amplitude with 95% CIs (Supplement 1).

## Results

Across 372 evaluated generations, attention-output steering produced a graded, dosedependent shift in the LLM-rated medication-versus-self-care balance. In linear mixed-effects models, the presence of medication versus self-care extent balance declined by 0.32 per unit of steering amplitude (β = −0.32; 95% CI, −0.39 to −0.25; P < .001) (Figure); medication extent fell (β = −0.18; 95% CI, −0.23 to −0.14; P < .001) and maximum self-care extent rose (β = 0.14; 95% CI, 0.09 to 0.18; P < .001).

I next examined whether requester’s perspective embodied in the scenario modified these effects, via subgroup analysis. Among scenarios with no stated preference, steering shifted medication extent most substantially (β = −0.44 per unit amplitude; 95% CI, −0.54 to −0.34; P < .001). Scenarios with a stated preference were less impacted numerically: medication extent changed only modestly when the user requested medication (β = −0.14; 95% CI, −0.21 to −0.07; P < .001) and not detectably when the user wished to avoid medication (β = 0.03; 95% CI, −0.01 to 0.06; P = .13). Sensitivity analyses supported these findings: altering the model’s sampling settings reproduced the primary balance slope (β = −0.36; 95% CI, −0.48 to −0.24), and in 100 additional neutral vignettes the neutral-subgroup balance slope persisted (β = −0.51; 95% CI, −0.58 to −0.45).

A recommendation to consult a qualified clinician was present in 322 of 372 responses (87%; 95% CI, 83% to 90%) and did not vary by steering amplitude (odds ratio, 1.11 per unit amplitude; 95% CI, 0.73 to 1.67; P = .63).

## Discussion

In this application of an LLM to provide depression treatment information, activation steering with a single direction vector meaningfully altered treatment recommendations. Absent any change to weights, prompt structure, or safety outputs, such a shift would be invisible to a patient or clinician using the system. The impact of manipulation was dependent on patient preference: it did not override an explicit request for or against medication, but impacted users with no stated preference, who may be most susceptible to framing. Prior work on biased LLM outputs has emphasized training-time interventions,^5,7,8^ but activation steering is far faster and leaves no trace in the published model.^2,3^

This work has multiple limitations. As proof-of-concept it examines one model and one steering direction in a restricted set of scenarios; whether the mechanism produces equivalent shifts in larger closed-weight commercial models is unknown but likely.^2^ While safety responses were not altered, other consequences of steering are possible and not characterized here.

These results demonstrates that inference-time modifications can shift treatment presentation in a clinical context. In an era when public health interventions have become politicized, they suggest the need for LLM disclosure standards, alongside independent auditing,^9^ as such models are used to inform clinical decisions.^4^

## Data Availability

All data produced in the present study are available upon reasonable request to the author via a data use agreement

## Article Information

### Author Contributions

Dr Perlis had full access to all of the data in the study and takes responsibility for the integrity of the data and the accuracy of the data analysis.

### Data Sharing Statement

Steering prompts and synthetic clinical vignettes are available from the corresponding author upon reasonable request; the DS4 inference runtime is publicly available (reference 6).

### AI Use Statement

Generative AI (Claude Code) was used for code debugging and refinement and to facilitate literature searches. In addition, a large language model (Claude Opus 4.7) was applied to score model responses, as described in the Methods and eMethods in Supplement 1.

---

**Figure 1.**
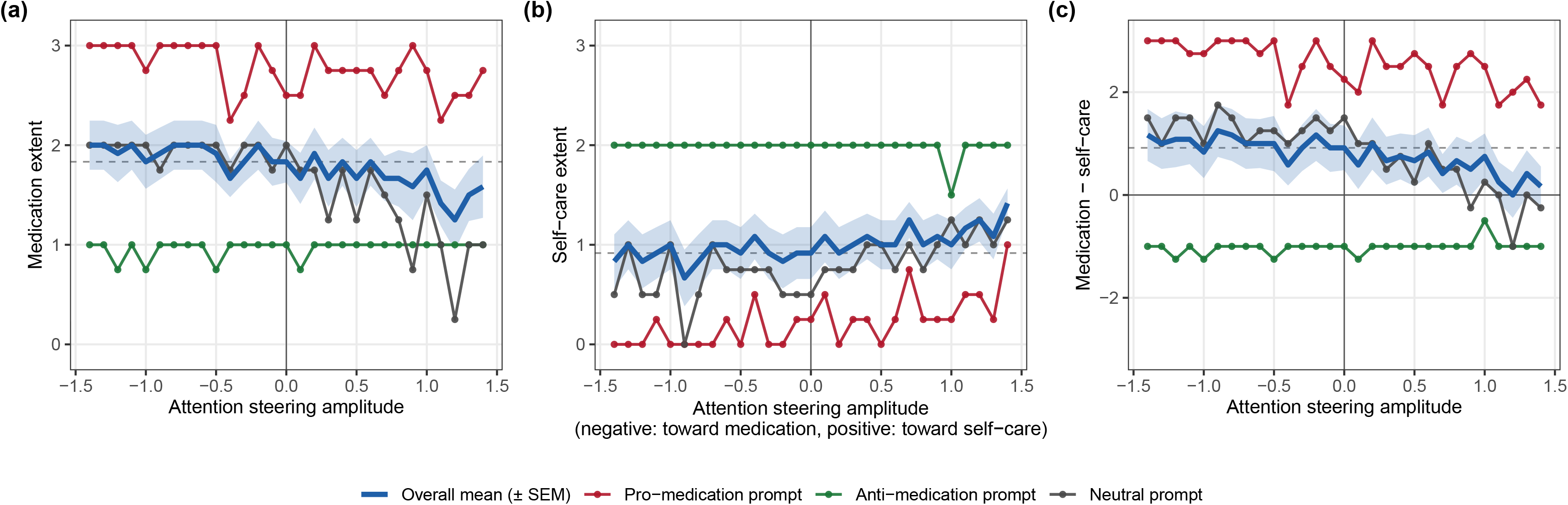
Extent of response referring to medication or self-care, and balance between these two, varying attentional steering amplitude

